# Prospective screening for monkeypox infection among men who have sex with men at an infectious diseases’ outpatient clinic in Copenhagen, Denmark

**DOI:** 10.1101/2025.01.06.24319307

**Authors:** Nuno Felipe Miranda Freire De Andrade, Mette Pinholt, Julie Maria Korsholm Mindedahl Nyamadi, Gitte Kronborg, Frederik Neess Engsig

## Abstract

Since 2022, over 102,000 cases of mpox clade IIb have been reported worldwide, primarily affecting men who have sex with men. Key risk factors include the use of HIV pre-exposure prophylaxis (PrEP) and HIV infection. In Denmark, the smallpox vaccine (Imvanex®) has been available to at-risk individuals since 2022. However, cases of reinfection with mpox have raised concerns about immunity, asymptomatic transmission.

Between August and October 2023, we screened 123 PrEP users for mpox at an outpatient clinic at Copenhagen University Hospital Hvidovre, Denmark. Mpox PCR testing was conducted using rectal swabs. Among the participants, 97% had received at least one dose of the mpox vaccine. No mpox cases were detected.

These findings suggest that, in a highly vaccinated high-risk population during a period of very low mpox prevalence in Denmark, asymptomatic mpox circulation is unlikely. The results support the current strategy of focusing mpox testing on symptomatic individuals and contact tracing during low-prevalence periods with high vaccination coverage.

## Main text

Globally, there have been 102,000 confirmed cases of mpox clade IIb (formerly known as monkeypox) and 220 deaths since January 2022.^1,2^ A significant rise in cases has been noted in countries where mpox transmission was not previously common. To date, 111 countries have reported cases, with Europe and the Americas experiencing the highest impact. In Denmark, 203 cases have been confirmed since May 2022, with 5 new cases between June and august 2024.^3^ The majority of cases have exhibited mild symptoms, primarily rash and fever.^4^

The outbreak is characterized by a notable shift in transmission patterns, with the virus spreading primarily through sexual contact among GBMSM (gay, bisexual, and other men who have sex with men).^5–7^ Notably, the use of HIV pre-exposure prophylaxis (PrEP) and HIV infection has been identified as significant risk factors for mpox infection with clade IIb. ^4,8^ Consequently, Danish authorities have offered smallpox vaccine (Imvanex®) to men who have sex with men since August 12, 2022, following World Health Organization guidelines.

In the aftermath of the outbreak more details of mpox infection in humans shows that mpox infection in patients with low CD4 cell count can result in fatal necrotizing infections and that re-infection is possible in both vaccinated and previously infected individuals.^9–13^ Re-infection implies that both natural immunity and vaccine-induced immunity are not fully protective.^14–17^ Furthermore, the re-infected individuals appear to have fewer and milder symptoms compared to persons with primary infection.^14^ Consequently, subclinical infections or maybe even asymptomatic carriage of mpox could theoretically be possible in a population.^18^ Indeed, asymptomatic carriage has been described in the beginning of the current outbreak but no screenings have been performed since in a setting where the majority of the high-risk population is presumed vaccinated.^19,20^ If mpox still is circulating in asymptomatic vaccinated persons, this could challenge current containment strategies.

To challenge this assumption further we screened Danish PrEP users at an infectious disease outpatient clinic for mpox. Between August 29, 2023, and October 13, 2023, men who have sex with men (MSM) who were screened for STI at the infectious disease outpatient clinic at Copenhagen University Hospital, Hvidovre, were offered additional screening for mpox. Mpox PCR testing was performed on rectal swaps and performed at the Department of Clinical Microbiology, Hvidovre, using the Novaplex MPXV Assay (Seegene, Seoul, South Korea) on the CFX Real-Time PCR system (BIO-RAD, California, USA). Chlamydia (*Chlamydia Trachomatis)* and gonorrhea (*Neisseria Gonorrhoeae*) testing was performed on swaps from throat, urine and rectum using the Aptima Combo 2 assay on the Panther system (Hologic, San Diego, USA). The screening was performed as a quality control study of STD screening and treatment.

A total of 123 MSM accepted screening for mpox. 118 (96%) were PrEP users and 5 (4%) were persons living with HIV. The median age of participants was 38.5 years. Ninety-seven (97%) study subjects had received at least one dose of the mpox vaccine, along with 73 (59%) of subjects being double vaccinated. Six (5%) persons tested positive for chlamydia and 11 (9%) for gonorrhea but none tested positive for mpox. This may be a due to a combination of 1) vaccination induced immunity; 2) isolation strategies following mpox diagnosis among high-risk persons; 3) a low general incidence of mpox in Denmark at the time of the screening; 4) a high level of awareness in the population at risk. The importance of vaccination has recently been emphasized by the ongoing outbreak in Australia, with more than 500 confirmed mpox cases in 2024 among whom nearly 50% were not vaccinated for mpox.^21^ No other studies have been published on screening for mpox in the post-vaccination era and we believe that this study contributes to the understanding of mpox transmission dynamics and Infection in a high-risk population in the post-vaccination era.

The incidence of chlamydia and gonorrhea found in the screened persons confirms that we screened a population at risk of being infected with mpox. Although studies have demonstrated a high sensitivity for mpox testing at the rectal mucosa, screening from other sites as well increased the sensitivity ^22,23^. This may be a limitation leading to missed cases. Still, none of the screened individuals were diagnosed with mpox subsequently.

The screening indicates that in a high-risk population, of whom the majority is vaccinated, in a period with low incidence of mpox, circulation of asymptomatic mpox rectally is not likely.

Various pro- and retrospective studies at different time points of the epidemic have demonstrated that screening asymptomatic individuals especially in periods with low incidence and high vaccination coverage is not indicated and our findings supports this strategy.^18,20,24–26^

## Data Availability

The nowcast data for all individual models are available at https://github.com/KITmetricslab/hospitalization-nowcast-hub. The code used to reproduce the results presented throughout this paper is available at https://github.com/avramaral/ensemble_learning.

https://github.com/avramaral/ensemble_learning

## Notes

### Competing Interest Statement

The authors have declared no competing interest.

### Funding Statement

This study did not receive any funding

### Author Declarations

Ethics committee/IRB of Region Hovedstaden; Hvidovre/Amager Hospital gave ethical approval for this work

## References

1. World Health Organization. (2024). 2022 Mpox (Monkeypox) Outbreak: Global Trends. (Accessed October 24, 2024). https://worldhealthorg.shinyapps.io/mpx_global/.

2. Ongoing Clade II Mpox Global Outbreak | Mpox | CDC. https://www.cdc.gov/mpox/outbreaks/2022/index-1.html.

3. Internationalt udbrud af mpox (abekopper)”; “Situationen i Danmark. “International outbreak of mpox.”; “Situation in Denmark”. https://www.ssi.dk/sygdomme-beredskab-og-forskning/sygdomsudbrud/2024_mpox.

4. Thornhill, J. P. et al. Human monkeypox virus infection in women and non-binary individuals during the 2022 outbreaks: a global case series. The Lancet 400, 1953–1965 (2022).

5. Laurenson-Schafer, H. et al. Description of the first global outbreak of mpox: an analysis of global surveillance data. Lancet Glob Health 11, e1012–e1023 (2023).

6. de Vries, H. J. et al. Mpox outbreak among men who have sex with men in Amsterdam and Rotterdam, the Netherlands: no evidence for undetected transmission prior to May 2022, a retrospective study. Eurosurveillance 28, 2200869 (2023).

7. Pan, D. et al. Transmission of monkeypox/mpox virus: A narrative review of environmental, viral, host, and population factors in relation to the 2022 international outbreak. J Med Virol 95, (2023).

8. Jia, L. et al. Cases of Monkeypox show highly-overlapping co-infection with HIV and syphilis. Front Public Health 11, 1276821 (2023).

9. Hazra, A. et al. Mpox in people with past infection or a complete vaccination course: a global case series. Lancet Infect Dis 24, 57–64 (2024).

10. Mitjà, O. et al. Mpox in people with advanced HIV infection: a global case series. The Lancet 401, 939–949 (2023).

11. Protopapas, K., Dimopoulou, D., Kalesis, N., Akinosoglou, K. & Moschopoulos, C. D. Mpox and Lessons Learned in the Light of the Recent Outbreak: A Narrative Review. Viruses 2024, Vol. 16, Page 1620 16, 1620 (2024).

12. Pinnetti, C. et al. Mpox as AIDS-defining event with a severe and protracted course: clinical, immunological, and virological implications. Lancet Infect Dis 24, e127–e135 (2024).

13. Corma-Gómez, A. et al. Long or complicated mpox in patients with uncontrolled HIV infection. J Med Virol 96, e29511 (2024).

14. Hazra, A. et al. Mpox in people with past infection or a complete vaccination course: a global case series. Lancet Infect Dis 24, 57–64 (2024).

15. Thy, M. et al. Breakthrough Infections after Postexposure Vaccination against Mpox. New England Journal of Medicine 387, 2477–2479 (2022).

16. Raccagni, A. R. et al. Two individuals with potential monkeypox virus reinfection. Lancet Infect Dis 23, 522–524 (2023).

17. Musumeci, S. et al. A Case of Mpox Reinfection. Clinical Infectious Diseases 77, 135–137 (2023).

18. Ferré, V. M. et al. Detection of Monkeypox Virus in Anorectal Swabs From Asymptomatic Men Who Have Sex With Men in a Sexually Transmitted Infection Screening Program in Paris, France. Ann Intern Med 175, 1491–1492 (2022).

19. Agustí, C. et al. Self-sampling monkeypox virus testing in high-risk populations, asymptomatic or with unrecognized Mpox, in Spain. Nature Communications 2023 14:1 14, 1–11 (2023).

20. De Baetselier, I. et al. Retrospective detection of asymptomatic monkeypox virus infections among male sexual health clinic attendees in Belgium. Nature Medicine 2022 28:11 28, 2288–2292 (2022).

21. NSW Mpox Surveillance Report (2024) Mpox NSW outbreak 2024 - week ending 26 October 2024. https://www.health.nsw.gov.au/Infectious/mpox/Pages/surveillance-reports.aspx.

22. Tarín-Vicente, E. J. et al. Clinical presentation and virological assessment of confirmed human monkeypox virus cases in Spain: a prospective observational cohort study. Lancet 400, 661 (2022).

23. Peiro-Mestres, A. et al. Frequent detection of monkeypox virus DNA in saliva, semen, and other clinical samples from 12 patients, Barcelona, Spain, May to June 2022. Eurosurveillance 27, (2022).

24. Grabmeier-Pfistershammer, K. et al. No evidence of asymptomatic monkeypox infection in a highly sexually active MSM population in Austria. HIV Med 25, 150–153 (2024).

25. Van Dijck, C. et al. Mpox screening in high-risk populations finds no asymptomatic cases. Lancet Microbe 4, e132–e133 (2023).

26. Satapathy, P. et al. Potentially Asymptomatic Infection of Monkeypox Virus: A Systematic Review and Meta-Analysis. Vaccines (Basel) 10, 2083 (2022).

